# Predicting first-onset depression in adolescents: Do general population models generalize to youth with ADHD?

**DOI:** 10.64898/2026.04.30.26351304

**Authors:** Shiqi Lu, Toby Wise, Deanna M. Barch, Georgina M. Hosang, Giorgia Michelini

## Abstract

**Background:** Most studies seeking to identify youth at increased risk for depression have developed prediction models using a limited set of risk factors in general population samples. It is unclear whether these models generalize to high-risk youth. Here, we developed machine learning algorithms to predict first-onset depression in youth from the general population and high-risk youth with attention-deficit/hyperactivity disorder (ADHD).

**Methods:** Participants were 4803 unrelated children from the ABCD study with no prior mood disorder and complete data at baseline (age 9-10 years) and 2-year follow-up. Support Vector Machine, Random Forest, and Elastic Net models were used to predict first-onsets from clinically-relevant risk factors spanning mental and physical health, cognitive, dispositional, interpersonal, and socio-environmental domains. Predictive performance was evaluated in the full sample and separately in participants with ADHD (N=584, 12.16%).

**Results:** Models trained on the full sample achieved good discriminative predictive power (area under the curve [AUC]=0.70 and accuracy=0.70-0.82). Predictors that replicated across models included earlier pubertal development, higher behavioral inhibition and aggression, and more time spent passively watching media content. In the ADHD subsample, model performance declined (AUC=0.46-0.61) and predictors only partly overlapped with those identified in the full sample.

**Conclusions:** Models effectively predicted depression in the general population but showed poor generalization to high-risk youth with ADHD, suggesting different risk factors in this group. These findings highlight that models trained in general population samples may not generalize to high-risk groups, pointing to the need for more tailored efforts to predict depression in youth at increased risk.

## Introduction

First onsets of depression are common in adolescence (∼20%)^1^, disrupting critical aspects of adolescent development and often leading to poor lifelong mental health and functional outcomes, including increased suicide rates^2^. There is growing interest in preventing depression before it emerges^3^, but previous attempts have largely relied on school-based, universal prevention programs that have failed to reduce depression incidence^4^. Adolescents with pre-existing mental health symptoms in particular show little improvement, or even negative effects^5^. Targeted approaches specifically focused on adolescents at high risk, rather than the adolescent general population, may be more beneficial and cost-effective^6^. However, our ability to identify those at highest risk to be prioritized for prevention remains limited, as research into risk factors for adolescent depression has scarcely been translated into effective early identification strategies^7,8^.

Previous prospective longitudinal studies of adolescent depression have established the role of clinical antecedents (e.g., sub-threshold depression), early pubertal development, parental mood disorder, cognitive and dispositional vulnerabilities (e.g., rumination), interpersonal factors (e.g., poor family relationships), and sociodemographic factors^9–12^. However, studies seeking to develop prediction models for adolescent depression based on candidate risk factors have generally investigated only a small number (<20) of risk factors using conventional statistical approaches^8,13^, which are limited in their ability to handle a large number of predictors and their complex interplay. Multi-informant and multimethod assessments of a wide range of risk factors are likely needed to capture the complex pathophysiology and heterogeneity of depression and aid screening/prevention strategies^7,8,10,14^.

Recently, researchers have leveraged machine learning to build data-driven prediction models of adolescent depression including a larger number of risk factors from various domains and accounting for potentially complex, non-linear relationships^9,15,16^. These machine learning studies seeking to predict adolescent depression diagnosis have yielded variable results, with studies on well-powered samples showing discrimination performance ranging from limited to good (area under the curve [AUC]=0.62-0.76)^16–19^. Yet, these models were developed from data collected ∼10-30 years ago in samples often characterized by limited socio-demographic diversity, not including assessments of potential risk factors that have been recognized more recently, such as screen time and cultural factors^9,20^. This limits their relevance and generalizability for future early identification efforts^21^.

Besides efforts to predict adolescent depression in the general population, developing prediction algorithms tailored to groups known for being particularly vulnerable can be an effective strategy for reducing depression incidence^6^. Adolescents with a history of attention-deficit/hyperactivity disorder (ADHD), representing 11.4% of the US youth population^22^, are one such group who should be prioritized for early identification and prevention. Youth with ADHD have at least twice as high risk for depression, earlier depression onsets (e.g., in early adolescence), longer and more chronic forms of depression greater suicide risk, and worse antidepressant response than youth without ADHD^23–25^. Yet, very few studies have investigated prospective risk factors for depression in this group. Besides the role of shared genetic factors^23^, available studies point to associations with interpersonal factors, such as peer rejection and victimization^26^, but have only included a limited number of predictors. Whether risk factors for depression in the general population are also predictive in this group remains unclear. Moreover, no study to date has leveraged multivariate approaches such as machine learning algorithms to examine whether general population models to predict depression (or internalizing/emotional disorders more generally) generalize to this high-risk population.

This study sought to address these gaps in the literature by developing machine learning models of first-onset depression in youth from the general population and high-risk youth with ADHD. We used longitudinal data from the Adolescent Brain Cognitive Development (ABCD) study^27,28^, the largest, most socio-demographically diverse, and most richly characterized longitudinal study of adolescents with multi-informant clinical assessments of depression diagnoses. Launched in 2016, assessments in this sample covered a comprehensive set of traditional risk factors for depression^10–12^, as well as emerging factors relevant to today’s adolescents from diverse backgrounds^9,20^. Recent studies aiming to predict depression in this sample have either included a limited number of risk factors or focused on predicting continuous depression ratings^9,17,29^. Here, we instead used the comprehensive range of risk factors available in ABCD to predict clinical diagnoses based on multi-informant symptom reports and functional impairment, which offer greater value for guiding early identification efforts.

Specifically, we first aimed to predict first-onset depression in the whole sample. Then, we tested the generalizability of the model developed in the full sample to adolescents with ADHD, in addition to developing a new model in this high-risk subsample. In addressing both aims, we took a largely data-driven approach, including a wide range of predictors that could be collected easily and at low cost in a wide range of settings (e.g., schools or primary care). Neuroimaging variables were excluded for this reason, and because they may increase overfitting^30^ and have shown limited ability to predict depression and other outcomes in this sample^9,31^. We broadly hypothesized that first onsets in the full sample would be predicted by a diverse range of risk factors, and that partly different risk factors would emerge in young people with ADHD, suggesting the need for a tailored approach.

## Methods

### Sample

The ABCD study recruited 11,875 children aged 9-10 years from 21 sites across the US, using probability sampling to ensure an unbiased representation of the sociodemographic diversity within the US population. Recruitment was primarily facilitated through private and public elementary schools^28,32^. At baseline, the sample approached the socio-demographic diversity of the US population^33^ (Table 1, Table S1). All study procedures were approved by Institutional Review Boards and formal written consent was provided by parents or legal guardians^34^.

**Table 1.** Demographic characteristics of participants with and without a first onset of depression.

|  | First onset<br>(N=477) |  | No onset<br>(N=4326) |  | First onset vs. No<br>onset |  |
| --- | --- | --- | --- | --- | --- | --- |
|  | Mean | SD | Mean | SD | t | p |
| Age at baseline | 9.52 | 0.5 | 9.46 | 0.51 | -2.91 | <0.01 |
| | N | % | N | % | $\chi^2$ | p |
| Sex |  |  |  |  | 8.47 | <0.01 |
| Male | 219 | 45.7 | 2287 | 52.84 |  |  |
| Female | 258 | 53.46 | 2038 | 47.09 |  |  |
| Family income |  |  |  |  | 19.46 | <0.01 |
| <\$50,000 | 127 | 26.62 | 890 | 20.57 | | |
| \$50,000-100,000 | 142 | 29.77 | 1171 | 27.07 | | |
| >\$100,000 | 172 | 36.06 | 2022 | 46.74 | | |
| Parental education |  |  |  |  | 2.72 | <0.01 |
| Neither parent holds a Bachelor's or higher degree | 214 | 44.87 | 1583 | 36.59 |  |  |
| One or both parents hold a Bachelor's or higher degree | 263 | 55.13 | 2743 | 63.41 |  |  |
| Race |  |  |  |  | 7.61 | 0.27 |
| White | 353 | 74 | 3296 | 76.19 |  |  |
| Black | 68 | 14.26 | 534 | 12.34 |  |  |
| Asian | 20 | 4.19 | 227 | 5.25 |  |  |
| American Indian/Alaska Native | 13 | 2.73 | 76 | 1.76 |  |  |
| Native Hawaiian/Pacific Islander | 0 | 0 | 8 | 0.18 |  |  |
| Other | 20 | 4.19 | 174 | 4.02 |  |  |
| Ethnicity |  |  |  |  | 2.77 | 0.25 |
| Hispanic or Latino/a/x | 394 | 82.6 | 3657 | 84.54 |  |  |
| Non-Hispanic or non-Latino/a/x | 79 | 16.56 | 652 | 15.07 |  |  |
| ADHD diagnosis |  |  |  |  | 55.58 | **<0.01** |
| With ADHD | 109 | 22.85 | 475 | 10.98 |  |  |
| Without ADHD | 368 | 77.15 | 3851 | 89.02 |  |  |

Abbreviations: ADHD = attention-deficit/hyperactivity disorder; SD=Standard Deviation.

The current study focused on risk factors measured at baseline and depression diagnoses between baseline (mean age=9.48, SD=0.51) and 2-year follow-up (N=10,908, 91.91% of the baseline cohort, mean age=11.60, SD=1.40), using data from the ABCD Release 5.1 (http://dx.doi.org/10.15154/z563-zd24). This is the first follow-up of the sample which assessed psychiatric diagnoses after baseline. Participants who completed 2-year follow-ups were more likely to be male, White, non-Hispanic, and from higher socio-economic backgrounds (Table S1). Of participants retained at follow-up, we further excluded from analyses participants with a history of depressive disorder or bipolar disorder at baseline (N=2,603), as our focus was on first-lifetime onsets (see Supplement 1 and Measures for further details). Additionally, we randomly included one child per family (excluding N=1,453) to maintain data independence and avoid overfitting in machine learning analyses. Finally, we excluded participants with incomplete data for baseline predictors, resulting in a final sample of 4,803 participants (see Figure S1 for a flowchart of participants’ exclusions).

### Measures

#### Baseline predictors

A comprehensive set of baseline predictors from baseline assessments was included using a largely hypothesis-free strategy, focused on capturing diverse aspects of risk for adolescent depression and including all measures that could be collected easily and at low cost in future early identification efforts, while excluding more specialized assessments (e.g., neuroimaging). In total, 114 predictors were included in the final analysis. For a full description of variables included and excluded from analyses, see Tables S2-3.

##### Mental health

Current and prior diagnoses of any mood, anxiety, eating, neurodevelopmental, and behavioral disorders were assessed using the computerized Kiddie Schedule of Affective Disorders and Schizophrenia for DSM-5 (KSADS-COMP), conducted separately with parents and children^35^. The KSADS was also used to identify participants with ADHD for our analyses on this high-risk subgroup. Following previous recommendations in this sample^36^, we considered participants to have a current or prior ADHD diagnosis if (1) they had at least 6 inattentive and/or hyperactive-impulsive symptoms and impairment across two or more settings, (2) they had an estimated IQ over 70 (based on the Wechsler Intelligence Scale for Children – Fifth Edition (WISC-V) Matrix Reasoning total scale score > 3), and (3) they did not meet criteria for schizophrenia or bipolar disorder.

Questionnaires further measured children’s emotional and behavioral problems, prodromal psychosis, early alcohol and substance use, as well as parents’ own mental health history and recent psychopathology.

##### Cognition and dispositional traits

Participants completed cognitive tasks assessing multiple cognitive skills, including sustained attention, memory, executive function, sensory function, delay discounting, verbal learning and memory, fluid intelligence and visuospatial reasoning. Parents also completed a range of questionnaires covering dispositional traits, including behavioral inhibition/behavioral activation and impulsivity.

##### Demographic characteristics, environment, culture and lifestyle

Age, gender, current school year, parents’ highest education, marital status, and language skills were included. Participants and parents also completed several assessments covering family characteristics (e.g., family conflict, cohesion, parental monitoring, and parental warmth), school environment (e.g., involvement, disengagement), neighborhood safety, cultural and family values, physical exercise habits, sleep, and screen time (passive watching media content, active gaming, social media use, and social engagement).

##### Physical health

Assessments of physical health included body mass index (BMI), waist circumference, pubertal development, and developmental and medical history (e.g., preterm birth, frequency of emergency room visits).

#### Follow-up depression diagnosis (outcome)

Diagnoses of depression between baseline and 2-year follow-up were assessed using the aforementioned K-SADS-COMP. We aggregated diagnoses of major depressive disorder, persistent depressive disorder (dysthymia), or unspecified depression (also known as depression not-otherwise specified), in line with previous studies of first-onset depression^8^. Unspecified depression was operationalized as a clinically significant depression (e.g., characterized by cardinal symptoms with suicidality, significant impairment, or need for treatment) that fell short of meeting the full criteria for major or persistent depressive disorder. We considered depression to be present if criteria on the KSADS-COMP interview were met based on either the child or the parent.

### Data analysis

#### Machine learning algorithms

Machine learning analyses were conducted in Python (Python Software Foundation, https://www.python.org/). All analysis code is publicly available at: https://osf.io/d3hv8/overview?view_only=3e4b38a67607414a9b18e5fc935c97cd

We used baseline risk factors as predictors of first-onset depression between baseline and two-year follow-up as the binary outcome. We first developed a prediction model for the general population by running analyses in the whole analysis sample, and then we ran analyses on participants with ADHD. A detailed description of machine learning analyses is provided in Supplement 2.

We selected three supervised machine learning algorithms that are widely used for classification/prediction, show robust performance, and can model linear and non-linear relationships^37^. Support Vector Machine (SVM) can effectively categorize individuals into groups within a statistical framework based on margins^37^, with linear or Radial Basis Function (RBF) kernels trying to separate different groups. Elastic Net models linear relationships and fits within the framework of regularized generalized linear models^38^. Random Forest is a decision tree-based algorithm that combines predictions from many single decision trees to solve regression or classification problems^37^. See Supplement 2 for a detailed description.

#### Preprocessing

Before conducting machine learning analyses, correlations between variables were assessed. No variables exhibited high collinearity, as all correlation coefficients were below 0.90. All categorical variables were dummy-coded, and continuous variables were standardized.

Variables with low variance were removed. In total, 114 variables included in the models. Participants were randomly split into training (80%) and testing (20%) sets, stratified by depression diagnosis to ensure similar representation across sets. To protect against overfitting and improve the performance of our models in independent datasets, regularization^39^ and hyperparameter tuning methods (grid search) were employed in the training set based on performance, as evaluated using 10-fold cross-validation^39^ (see Supplement 2 for details).

Participants with first-onset depression were expected to account for a small portion of the total sample, making the positive class (depression present) underrepresented^9,29,40^. To address this imbalance, we incorporated class weights into the machine learning models to ensure adequate learning from the minority class (Supplement 3).

#### Evaluation metrics and feature importance

We evaluated model performance using accuracy, area under the curve (AUC) of the Receiver Operating Characteristics (ROC) curve, precision, recall, and F1 score in the independent test set. Our evaluation mainly relied on AUC, allowing us to appropriately capture model performance beyond overall accuracy in our imbalanced dataset i.e., more unaffected than affected participants), in line with gold-standard procedures^40–43^. The confusion metrics with a detailed breakdown of the model performance are shown in Table S3.

To compare the importance of individual predictors, we employed the SHapley Additive exPlanations (SHAP) Python package, which provides a unified framework for interpreting model predictions and identifying the most influential predictors in the model^44^. This allowed us to identify risk factors with high importance across different models. Despite inherent uncertainties in single machine learning predictions, identifying the same risk factors consistently across different algorithms points to reliable predictors^45^.

#### Permutation tests

To evaluate the significance of our model learning results, we carried out permutation tests for each model. This was achieved by shuffling the outcome variable, making predictions on the randomized outcome, and determining how likely our observed level of predictive accuracy was based on this empirical null distribution.

#### Prediction with the top 10 important risk factors

As an additional analysis, we also constructed streamlined versions of each algorithm, only including the top 10 most important variables. This allowed us to assess whether a reduced set of risk factors could still achieve robust prediction performance.

#### Cross-model evaluation

To assess whether the model developed in the full (general population) sample generalizes to participants with ADHD, we conducted cross-model evaluation. Specifically, the prediction model trained on the same training set used in the afore-mentioned analyses was tested on a subset of the original test set that only included participants with ADHD. This approach enabled us to determine whether a model developed in the general population retained predictive utility within higher-risk adolescents with ADHD.

#### ADHD model evaluation

Additionally, to test prediction performance in the high-risk ADHD group, we developed machine learning models only using participants with ADHD, following the same procedure outlined in the full sample. In these models, participants with ADHD were randomly assigned to either the training (80%) or testing (20%) set. This step was to compare the performance of this ADHD model with the performance of the model developed in the full sample, and examine if the risk profile is comparable or different in youth with ADHD.

## Results

Over the two-year follow-up period, N=477 participants (9.93% of the analysis sample) experienced a first-onset depression (46.6% major depressive disorder, 2.2% persistent depressive disorder, and 51.2% unspecified depression). Participants with and without a first onset did not significantly differ on race/ethnicity, but those with depression were slightly older and more likely to be female and come from families with lower parental educational attainment and household income (Table 1).

A total of N=584 participants (12.16% of the final sample) met criteria for present or past ADHD. Although this is slightly higher than US population estimates^22^, this group likely represents individuals meeting ADHD criteria or with clinically significant and impairing subthreshold presentations, showing elevated risk for poor mental health outcomes^23^. Indeed, youth with ADHD were more than twice as likely to have first-onset depression (18.66%) than those without ADHD (8.72%) (ꭓ^2^=56.69, p<0.01).

### Model performance in the whole sample

All three machine learning models predicting depression onset in the full sample achieved AUC=0.70 in the independent test set, demonstrating good discriminative capability across different algorithms (Table 2 and Figure 1). Accuracy was also consistently high (0.70-0.82), particularly for the Random Forest model, indicating good overall classification performance. Earlier puberty, higher behavioral inhibition and aggression, more time spent passively watching media content, and lower working memory emerged consistently as important predictors in SHAP analyses across all three machine learning algorithms (Table 3, Figure 2). Additional risk factors emerging across two of the three algorithms spanned higher prodromal psychosis, anxious/depressed and somatic problems, sleep problems, BMI and older age.

**Figure 1.**
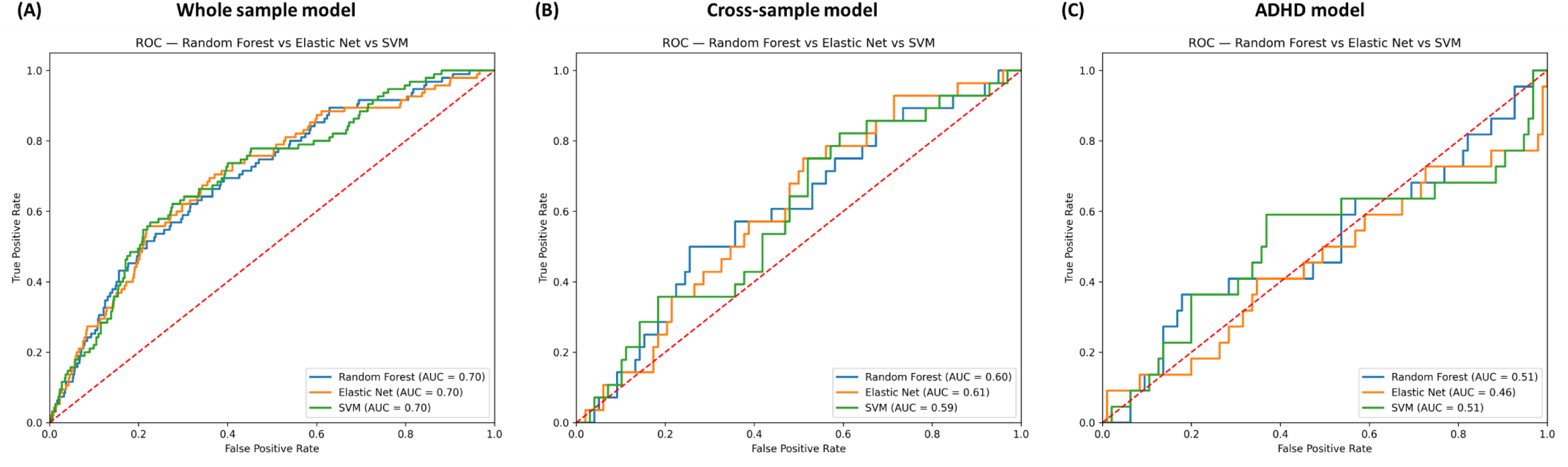
The ROC curves for (A) whole sample model, (B) cross-sample model (applying the models developed in the whole sample to the ADHD subsample), and (C) ADHD subsample model. Abbreviations: ADHD = attention-deficit/hyperactivity disorder, AUC = area under the curve; ROC = Receiver Operative Characteristics; SVM = Support Vector Machine.

**Figure 2.**
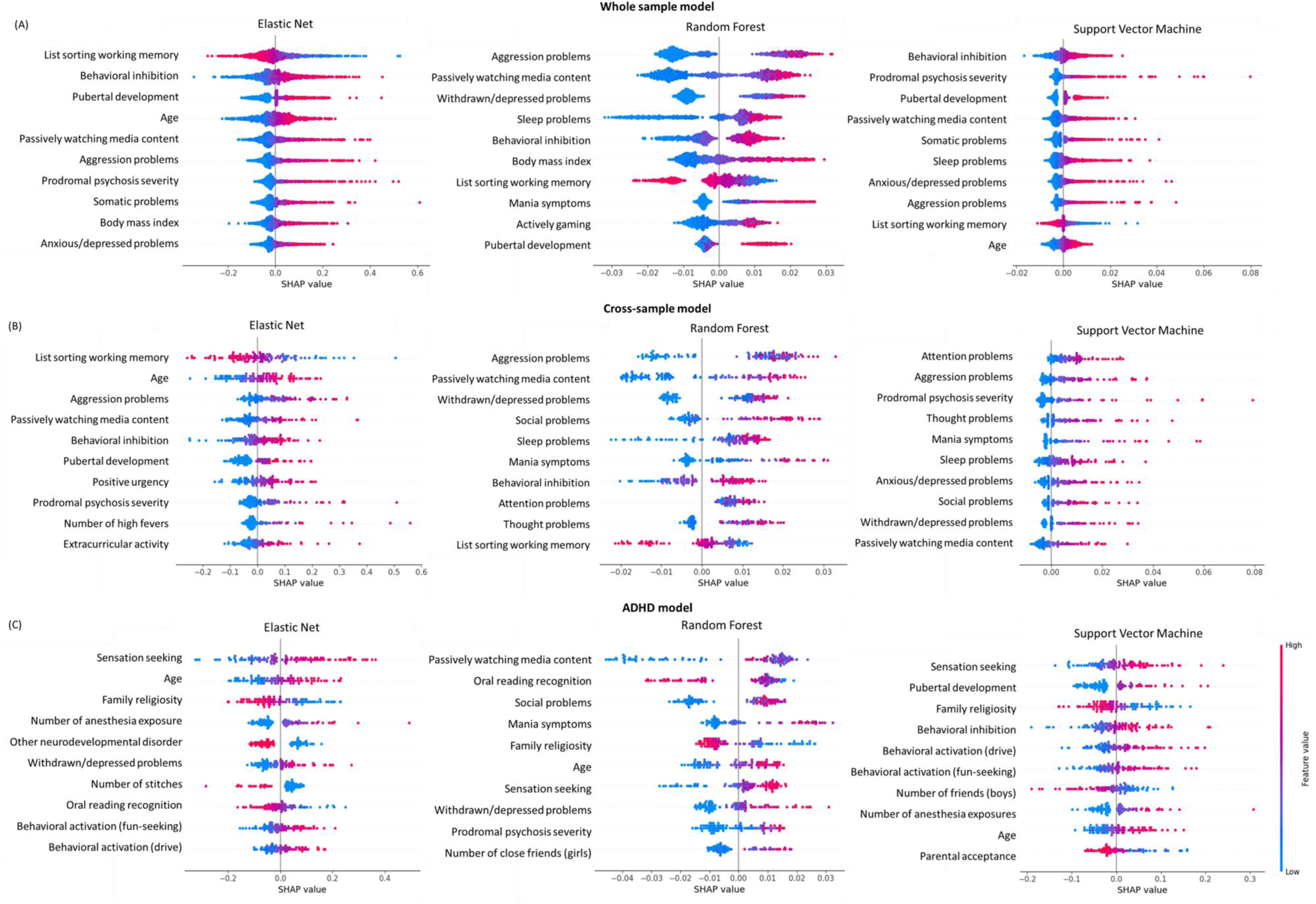
The top 10 most important features influencing machine learning predictions of first-onset depression across three models: (A) whole sample model, (B) cross-sample model (applying the models developed in the whole sample to the ADHD subsample), and (C) ADHD subsample model. Notes: These summary plots show the relationship between risk factors and model prediction. The y-axis includes feature importance ranked from most important, so the feature ranked first is the most important risk factor influencing first-onsets in that specific algorithm. The values on the x-axis represent the SHapley Additive exPlanations (SHAP) values, where values on the left-hand side indicate a lower likelihood to be predicted as belonging to the first-onset group, whereas values on the right-hand side indicate a higher likelihood. Each dot on the plot refers to the SHAP value for a participant on a given feature, where red dots indicate high feature scores, and blue dots indicate low scores. For example, in the Elastic Net model trained on the whole sample, list sorting working memory is ranked as the most important predictor, where lower scores indicate worse working memory. For this variable, lower scores (i.e., red dots) on the right-hand side of the SHAP plot suggest that a participant with worse working memory is more likely to be predicted as having a first onset, whereas a participant with better working memory (i.e., blue dots on the left hand-side) is more likely to be predicted as no onset.

**Table 2.**
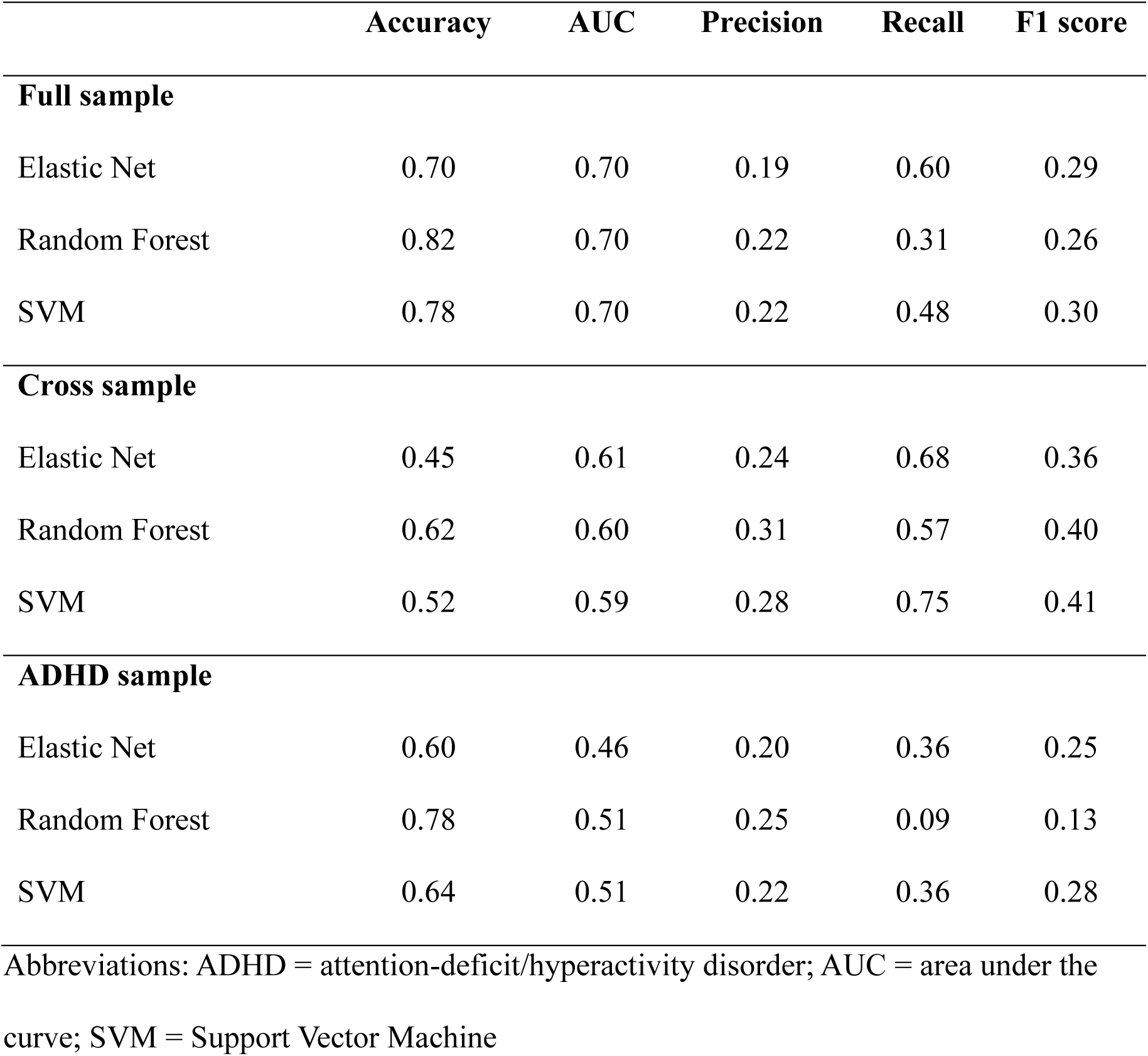
Performance metrics of all machine learning models.

|  | **Accuracy** | **AUC** | **Precision** | **Recall** | **F1 score** |
| --- | --- | --- | --- | --- | --- |
| **Full sample** |  |  |  |  |  |
| Elastic Net | 0.70 | 0.70 | 0.19 | 0.60 | 0.29 |
| Random Forest | 0.82 | 0.70 | 0.22 | 0.31 | 0.26 |
| SVM | 0.78 | 0.70 | 0.22 | 0.48 | 0.30 |
| **Cross sample** |  |  |  |  |  |
| Elastic Net | 0.45 | 0.61 | 0.24 | 0.68 | 0.36 |
| Random Forest | 0.62 | 0.60 | 0.31 | 0.57 | 0.40 |
| SVM | 0.52 | 0.59 | 0.28 | 0.75 | 0.41 |
| **ADHD sample** |  |  |  |  |  |
| Elastic Net | 0.60 | 0.46 | 0.20 | 0.36 | 0.25 |
| Random Forest | 0.78 | 0.51 | 0.25 | 0.09 | 0.13 |
| SVM | 0.64 | 0.51 | 0.22 | 0.36 | 0.28 |

Abbreviations: ADHD = attention-deficit/hyperactivity disorder; AUC = area under the curve; SVM = Support Vector Machine

**Table 3.**
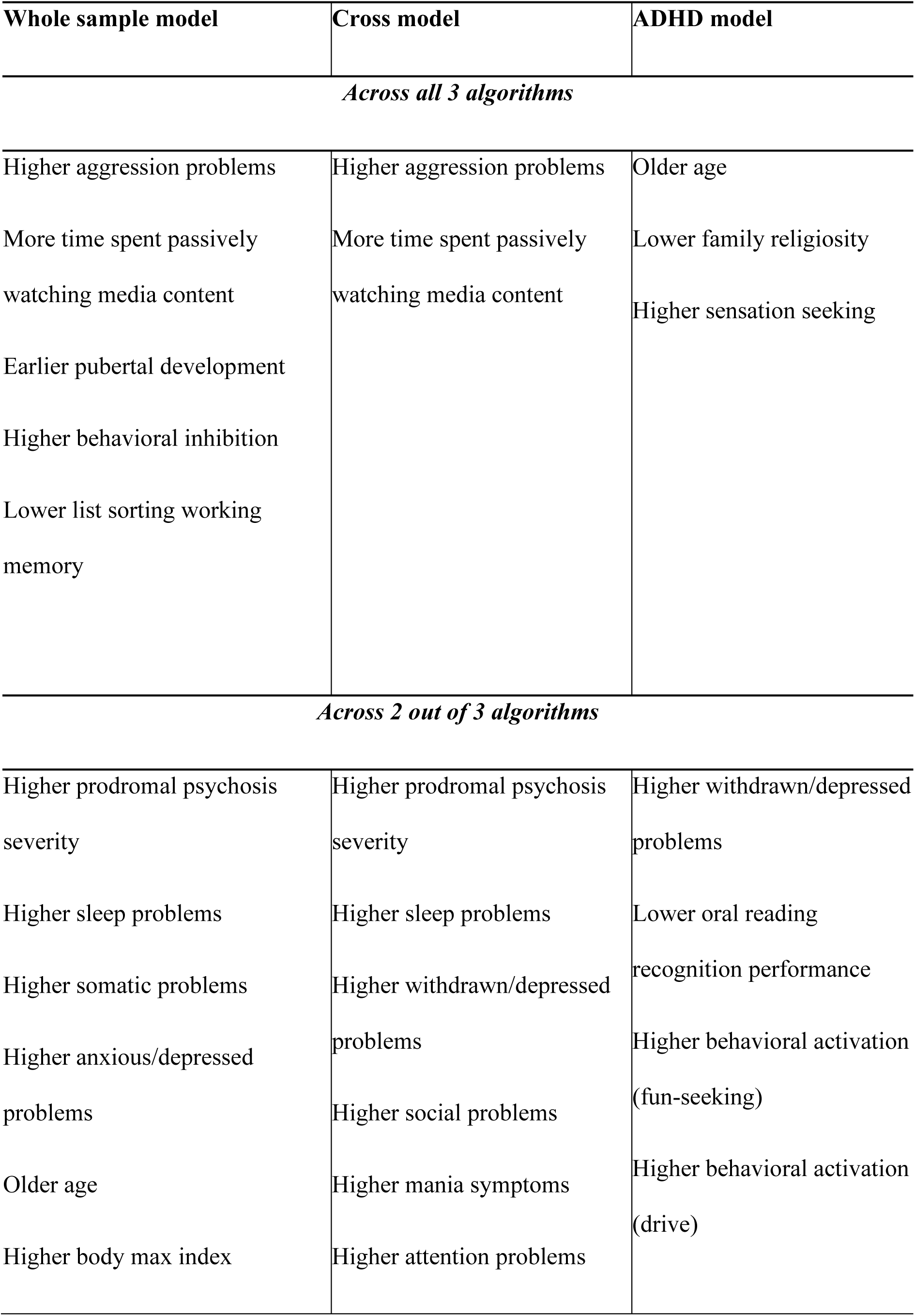
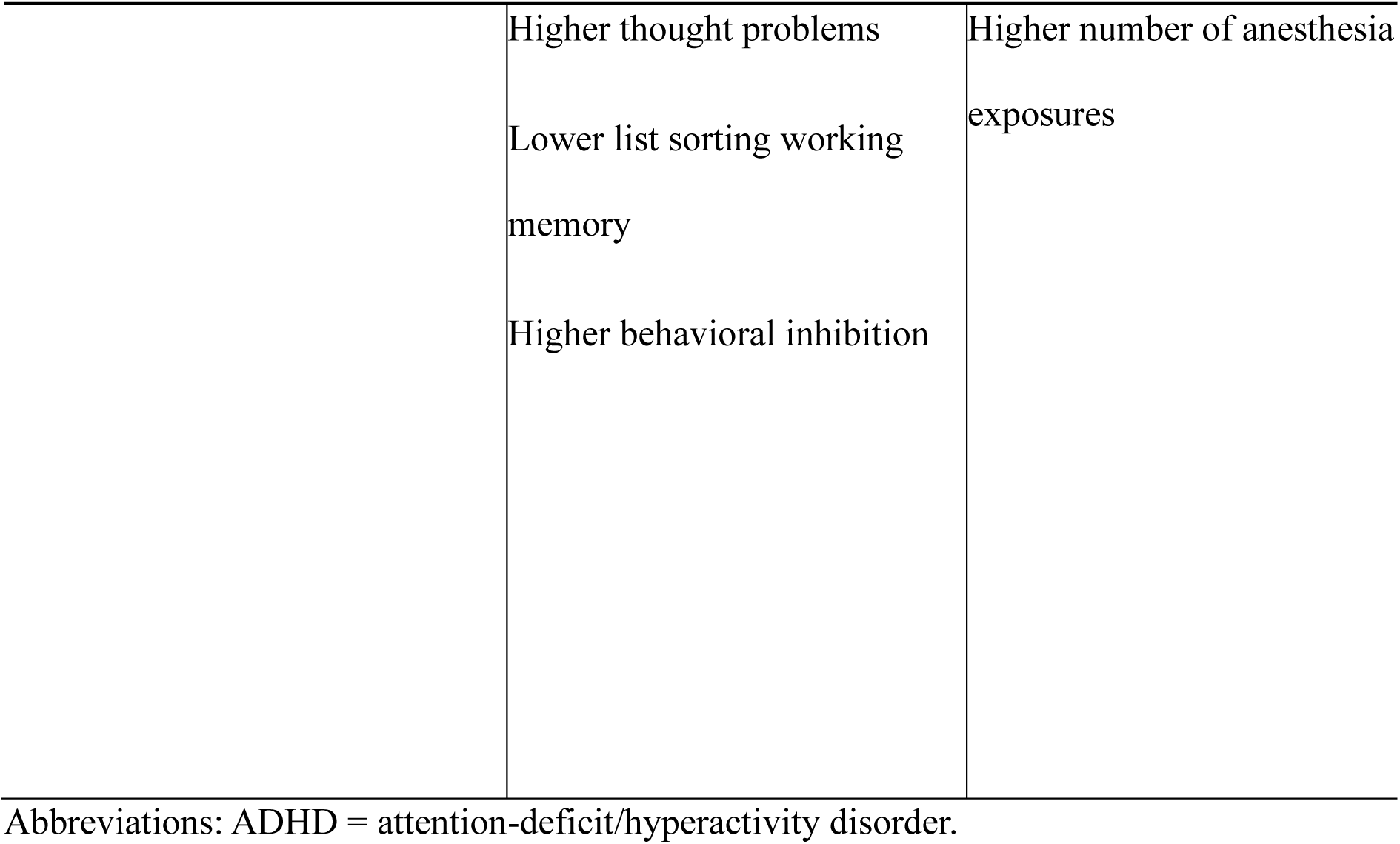
Important risk factors generated consistently across machine learning models.

When models were trained using only the top 10 most important predictors identified by the SHAP analyses (Figure S2), performance remained comparable for SVM and Elastic Net (both AUC=0.69), but dropped slightly for Random Forest (AUC=0.65).

### Prediction of depression in youth with ADHD using the general population model

In the cross-model evaluation, prediction models were trained on the whole sample training set using an identical set of predictors (excluding only ADHD diagnosis) and then tested on an independent ADHD subsample. This enables us to assess whether predictors of first onsets identified in the general population could generalize to youth with ADHD. Models achieved AUC=0.59-0.61, suggesting poor discriminative power (Table 2). SHAP analysis revealed that important predictors emerging consistently across the two or three algorithms included some of the predictors that showed consistency in the whole sample (e.g., aggression, passively watching media content, sleep problems, behavioral inhibition, prodromal psychosis, working memory), but not all (e.g., attention, thought, withdrawn/depressed and social problems, and mania symptoms emerged in cross-model analyses, but not in whole sample models) (Table 3, Figure 2).

### Prediction of depression in youth with ADHD using the tailored ADHD model

In models developed and validated within the ADHD subsample, the discriminative performance was poor (AUC=0.46-0.51) (Table 2). SHAP analysis showed that most of the important predictors replicating across algorithms did not overlap with those identified in the whole sample or cross model analyses (Table 3, Figure 2). Specifically, higher sensation seeking, behavioral activation (drive and fun seeking) and number of anesthesia exposures, and lower scores on family religiosity and oral reading performance emerged uniquely in the ADHD subsample across two or three of the machine learning algorithms, but did not emerge as top predictors in any of the whole sample or cross model analyses (Figure 2).

## Discussion

This study applied machine learning algorithms to predict diagnoses of first-onset depression in a large, population-based cohort of young adolescents, using comprehensive multi-informant assessments spanning mental and physical health, cognitive, dispositional, interpersonal, and socio-environmental domains. We found good predictive performance in the general population (whole sample model), indicating the feasibility of using prediction algorithms to identify youth at elevated risk for developing clinical depression. Our data-driven approach robustly identified predictors consistent with prior literature, such as earlier puberty and behavioral inhibition, as well as novel predictors not previously considered in prediction models, such as time spent passively watching media content. However, predictive accuracy declined substantially when predicting first-onsets in adolescents with ADHD, both when applying models developed in the whole sample and new models developed in the ADHD subgroup. Predictors emerging as important in the whole sample only partly overlapped with those in the ADHD subsample, pointing to potentially distinct risk profiles in youth with ADHD. Together, these findings highlight the importance of targeted approaches to prediction and early intervention/prevention of depression tailored to individual risk profiles, both in the general population and in high-risk groups.

The first major contribution of our study is the development of prediction models for first-onset adolescent depression in the general population, achieving an AUC of 0.70. This AUC has potential clinical value, as it reflects acceptable performance for scalable, low-cost screening strategies in population settings (e.g., schools)^18^. To put these values in context, comparable AUC scores have been reported for validated prognostic/prediction models currently used in clinical settings for other mental health conditions, such as psychosis^41^, as well as physical health conditions, including breast cancer and cardiovascular diseases^42,43^. While broad assessments of all risk factors (>100) included in our models would likely not be feasible in clinical or community settings, we found similarly acceptable performance in the whole sample when predicting depression only using the top 10 predictors identified in our models. Our findings extend previous studies based on large and diverse samples such as ABCD to predict depression, which used a small set of predictors or relied on continuous symptom ratings not accounting for multiple informants and functional impairment^9,15,29^. By considering a comprehensive set of predictors to forecast first-lifetime onset of depressive disorder, our study takes into account the multidimensional nature of depression risk and aligns closely with real-life scenarios that require identification of youth at highest risk of developing depression, enhancing the translation value of our prediction models.

Our prediction model identified a set of meaningful predictors of first-onset depression in the general population, which emerged consistently across all or most of our machine learning algorithms. Several of these predictors clustered within broader domains, offering insights into risk processes. Youth at greater risk for first-onset depression tended to show worse baseline mental health, including higher levels of prodromal psychosis, aggression, and anxiety/depression symptoms. This confirms the role of transdiagnostic mental distress in the emergence of depression^9^. These factors emerged alongside cognitive and temperamental vulnerabilities (worse working memory, and higher behavioral inhibition) that are broadly associated with mental health problems^7^. Besides mental health, the emergence of earlier pubertal development and higher BMI and somatic complaints as important predictors highlights the need to consider physical health factors in depression prediction models. Finally, our findings indicate the role of lifestyle factors, as youth spending more time passively watching media content and showing higher sleep problems (often associated with greater screen use) were more likely to have a first onset. Taken together, these findings highlight the role of emerging mental health problems and cognitive-dispositional traits, but also physical health and disengagement from social life as key predictors of first-onset depression in the general population^7,11,15,20^.

While a number of these risk factors, such as subthreshold depression, antecedent anxiety and cognitive-temperamental factors, have been included in previous models to predict adolescent first-onsets of depression^8,15^, our machine learning models also revealed novel predictors that have not been considered previously in these models, such as passive media use and pubertal development. Interestingly, although our analyses include several measures capturing interpersonal factors and adversity, these did not emerge among the most predictive risk factors, in contrast with their inclusion in previous (often hypothesis-driven) prediction models^15^. This might be due to the regularization step in machine learning algorithms, whereby the model weights of correlated measures are adjusted based on the impact of each measure on model performance^39^. Lower importance of interpersonal or adversity-related predictors may be explained by the emergence in our models of early puberty and aggression as prominent risk factors, which have been linked with exposure to threat-related adversity or unsupportive environments. Thus, they may thus represent more proximal risk factors for depression, resulting in greater predictive power in prediction models^10,12^.

The second major contribution of our study is that we sought to predict depression from a comprehensive set of risk factors in a particularly high-risk group, adolescents with ADHD. For the first time, we tested the ability of models developed through a general population approach (using our whole sample) to identify depression in this high-risk group. The predictive performance dropped substantially, with AUC=0.59-0.61, indicating poor discriminatory ability, only slightly better than chance. This low predictive performance in youth with ADHD highlights the limited generalizability of models trained on a large, general population sample to specific subgroups^46^. These findings indicate differences in risk profiles across young people from the general population and those with ADHD. As such, predicting depression in ADHD groups requires a more tailored approach.

We next adopted such an approach in developing and testing a model specifically for participants with ADHD. Although these analyses involved a relatively small sample (N=584), likely impacting predictive perfomance^40^, we found a limited overlap in the predictors emerging as important across analyses in the whole sample and the ADHD subgroup. Several predictors, such as cultural factors (higher family religiosity) and reward-seeking behaviors (higher drive, and fun and sensation seeking), emerged uniquely in youth with ADHD. Interestingly, the latter finding in youth with ADHD shows the opposite pattern to studies in the general population^47^, where *lower* reward sensitivity has been associated with depression risk^8^. This highlights that prediction models and risk profiles in the general population may not generalize to youth with ADHD. Overall, our dual approach to depression prediction in young people with ADHD underscores the need for more tailored prediction approaches in high-risk groups. Future studies should extend this approach to larger samples of young people with ADHD and potentially other neurodevelopmental conditions at increased depression risk.

Looking ahead, our prediction model in the whole sample may show practical utility for identifying youth from the general population at elevated risk for depressive disorder in early adolescence. Conversely, applying this model to high-risk youth with ADHD is unlikely to correctly identify those at increased depression risk. Future studies will need to externally validate this model in independent general population samples and test whether depression can be prevented in general population youth identified as at-risk through this model. Of note, the predictors identified in our models in the whole sample, such as early puberty and increased passive screen use, should be interpreted as important “warning signs” preceding the onset of depression, which may be easier to notice by clinicians, family members, and teachers than youth’s internal emotional difficulties. Yet, these risk factors do not necessarily represent intervention targets to prevent depression onsets in at-risk youth, as future work is needed to establish whether they are causally associated with future depression or instead represent indicators of escalating depression risk.

The strengths of this study include the use of a large socio-economically and racially/ethnically diverse sample, sophisticated analytic approach across general population and high-risk groups, and comprehensive assessments of clinically-relevant risk factors. Nevertheless, the following limitations should be considered. First, our study relied only on two timepoints spanning early adolescence, as the transition to adolescence is a period of particularly high risk for the development of depression, especially in youth with ADHD^23^. Future studies should extend this work to establish if findings generalize to first onsets in later adolescence. Second, the inclusion of predictors measured only at baseline prevented us from modeling the dynamic nature of risk over time. As emerging data suggest that depression first-onsets are predicted by persistently elevated levels of risk^14^, future studies should investigate the added predictive value of considering risk across multiple time points, particularly as new waves of data become available. Third, while our models achieved good AUCs of 0.70, it is possible that the inclusion of additional predictors requiring more specialized assessments (such as genetic or neuroimaging data) could further enhance model performance. Such predictors were not included in our study, as we focused on clinically-relevant and readily available predictors that could be collected easily in community or primary care settings. Future studies should evaluate the added predictive value of additional markers in combination with their feasibility and acceptability in real-world settings, including cost-benefit analyses. Finally, our ADHD subsample was relatively small for machine learning analyses. While focusing on individuals with ADHD from an epidemiological sample addresses typical limitations of clinically-referred samples, future studies should include larger ADHD samples and potentially more specialized assessments to achieve more robust prediction.

In conclusion, using a wide range of risk factors across multiple domains, we developed prediction models capable of predicting first onsets of adolescent depression in a socio-demographically diverse sample. Findings in the general population highlight the importance of considering a set of vulnerabilities spanning prior mental health, physical health, cognitive-dispositional traits and lifestyle factors in prediction models to identify youth at increased risk, who should be prioritized for prevention strategies. Additionally, prediction models developed in general population samples may not generalize to high-risk populations such as young people with ADHD, likely due to different risk profiles. Taken together, our data support the value of tailored approaches to early identification and prevention of mental health problems in young people, highlighting the necessity of developing specific screening tools that reflect the clinical profile of high-risk groups.

## Data Availability

Researchers can access the ABCD data via their data sharing procedures. The code for analyzing the data is available in OSF.

https://osf.io/d3hv8/overview

## Acknowledgements

The authors wish to thank all the families who participate in the ABCD Study and all of the ABCD Study staff who make this work possible. Data used in the preparation of this article were obtained from the Adolescent Brain Cognitive Development (ABCD) Study (https://abcdstudy.org), held in the National Institute of Mental Health Data Archive (NDA).

The ABCD Study is supported by the National Institutes of Health (NIH) and additional federal partners under awards numbers U01DA041048, U01DA050989, U01DA051016, U01DA041022, U01DA051018, U01DA051037, U01DA050987, U01DA041174, U01DA041106, U01DA041117, U01DA041028, U01DA041134, U01DA050988, U01DA051039, U01DA041156, U01DA041025, U01DA041120, U01DA051038, U01DA041148, U01DA041093, U01DA041089, U24DA041123, U24DA041147. A full list of supporters is available at https://abcdstudy.org/nih-collaborators. A listing of participating sites and a complete listing of the study investigators can be found at https://abcdstudy.org/principal-investigators.html. ABCD consortium investigators designed and implemented the study and/or provided data but did not necessarily participate in analysis or writing of this report. This manuscript reflects the views of the authors and may not reflect the opinions or views of the NIH or ABCD consortium investigators. The ABCD data repository grows and changes over time. The ABCD data used in this report came from DOI: <u>10.15154/z563-zd24</u>. SL was supported by a Chinese Scholarship Council PhD studentship. TW was supported by a Wellcome Trust Career Development Award (225945/Z/22/Z). GM was part-funded by a Klingenstein Third Generation Foundation Fellowship (20212999).

## Presentation information

Preliminary findings from this study were presented as a poster at the 2024 European Network of Hyperkinetic Disorders (EUNETHYDIS) meeting in Cagliari, Italy, in October 2024, and at the 2025 British Association of Psychopharmacology (BAP) meeting in Manchester, UK, in July 2025.

## CRediT authorship contribution statement

SL: Conceptualization, Methodology, Formal analysis, Data Curation, Investigation, Visualization, Writing – original draft, Writing – review & editing. TW: Methodology, Data Curation, Supervision, Writing – review & editing. DMB: Data Curation, Investigation, Funding acquisition, Writing – review & editing. GMH: Methodology, Supervision, Writing – review & editing. GM: Conceptualization, Methodology, Investigation, Supervision, Funding acquisition, Writing – original draft, Writing – review & editing.

