## Supplementary material for "Predicting first-onset depression in adolescents: Do general population models generalize to youth with ADHD?": SUPPLEMENTARY MATERIALS_feedback_final.docx

**Supplement 1**. **Exclusion of mood disorder at baseline**

Our analyses focused on participants without a prior history of mood disorder, including major depressive disorder, persistent depressive disorder (dysthymia) and unspecified depression, since our focus was on prediction of first lifetime onset of depression. According to the ABCD release notes (https://nda.nih.gov/study.html?id=2313), the KSADS computer diagnostic algorithm may overestimate symptoms of mania, potentially leading to inflated rates of bipolar disorder diagnoses. To address this, requiring at least one episode of past or present depression diagnoses (e.g., MDD, dysthymia, other specified depression) in addition to mania has been recommended. Additionally, parent reports are regarded as superior for identifying bipolar disorder in child and adolescent populations^1^. Therefore, in our study, we combined child and parent reports of depression at baseline but exclusively relied on parent reports for bipolar disorder diagnosis to exclude participants with baseline mood disorders.

**Table S1.** Description of participants' socio-demographic characteristics.

|  |  | **Retained at follow-up (N=10908)** | | **Lost at follow-up (N=960)** | | **Retained vs. Lost** | | **Included in the machine learning models (N=4803)** | | **Not included in the final model**  **(N=7065)** | | **Included vs. not included in model** | |
| --- | --- | --- | --- | --- | --- | --- | --- | --- | --- | --- | --- | --- | --- |
|  |  | **Mean** | **SD** | **Mean** | **SD** | **t** | **p** | **Mean** | **SD** | **Mean** | **SD** | **t** | **p** |
|  | Age at baseline | 9.48 | 0.51 | 9.47 | 0.52 | -0.38 | 0.70 | 9.47 | 0.51 | 9.49 | 0.51 | 1.57 | 0.12 |
|  |  | **N** | **%** | **N** | **%** | $\boldsymbol{\chi}^{\mathbf{2}}$ | **p** | **N** | **%** | **N** | **%** | $\boldsymbol{\chi}^{\boldsymbol{2}}$ | **p** |
|  | Sex |  |  |  |  | 5.54 | **0.02** |  |  |  |  | 0.07 | 0.97 |
|  | *Male* | 5718 | 52.50 | 463 | 48.53 |  |  | 2504 | 52.21 | 3677 | 54.56 |  |  |
|  | *Female* | 5173 | 47.50 | 491 | 51.47 |  |  | 2292 | 47.79 | 3372 | 50.04 |  |  |
|  | Family income |  |  |  |  | 132.62 | **<0.01** |  |  |  |  | 214.61 | **<0.01** |
|  | *<$50,000* | 2837 | 28.27 | 385 | 47.18 |  |  | 1017 | 22.48 | 2205 | 34.85 |  |  |
|  | *$50,000-100,000* | 2874 | 28.64 | 194 | 23.77 |  |  | 1313 | 29.02 | 1755 | 27.74 |  |  |
|  | *>$100,000* | 4324 | 43.09 | 237 | 29.05 |  |  | 2194 | 48.50 | 2367 | 37.41 |  |  |
|  | Race |  |  |  |  | 91.89 | **<0.01** |  |  |  | | 111.30 | **<0.01** |
|  | *White* | 7791 | 73.00 | 547 | 56.98 |  |  | 3649 | 76.20 | 4689 | 68.83 |  |  |
|  | *Black* | 1596 | 14.95 | 230 | 23.96 |  |  | 602 | 12.57 | 1224 | 17.97 |  |  |
|  | *Asian* | 488 | 4.57 | 49 | 5.10 |  |  | 247 | 5.16 | 290 | 4.27 |  |  |
|  | *American Indian/Alaska* | 269 | 2.52 | 37 | 3.85 |  |  | 89 | 1.86 | 217 | 3.19 |  |  |
|  | *Native Hawaiian/Pacific* | 31 | 0.29 | 2 | 0.21 |  |  | 8 | 0.16 | 25 | 0.37 |  |  |
|  | *Other* | 498 | 4.67 | 63 | 6.56 |  |  | 194 | 4.05 | 367 | 5.39 |  |  |
| Ethnicity | |  |  |  |  | 33.31 | **<0.01** |  |  |  |  | 28.99 | **<0.01** |
| *Hispanic or latino/a* | | 2148 | 19.93 | 262 | 27.93 |  |  | 860 | 18.12 | 1550 | 22.24 |  |  |
| *Non-hispanic or non-latino/a* | | 8629 | 80.07 | 676 | 72.07 |  |  | 3885 | 81.88 | 5420 | 77.76 |  |  |
|  | Parental education |  |  |  |  | 147.54 | **<0.01** |  |  |  |  | 320.92 | **<0.01** |
|  | *Neither parent with a Bachelor's or higher degree* | 2753 | 31.53 | 339 | 50.60 |  |  | 1104 | 26.86 | 1988 | 37.57 |  |  |
|  | *One or both parents with a Bachelor's or higher degree* | 5978 | 68.47 | 331 | 49.40 |  |  | 3006 | 73.14 | 3303 | 62.43 |  |  |
|  | ADHD diagnosis |  |  |  |  | 0.25 | **0.03** |  |  |  |  | 4.30 | **0.04** |
|  | *With ADHD* | 1373 | 12.78 | 137 | 14.62 |  |  | 584 | 12.16 | 926 | 13.47 |  |  |
|  | *Without ADHD* | 9369 | 87.22 | 800 | 85.38 |  |  | 4219 | 87.84 | 5950 | 86.53 |  |  |

Abbreviations: ADHD=Attention-Deficit/Hyperactivity Disorder, SD=Standard Deviation

Notes: Participants included and excluded from analyses are shown in Figure S1 (final analytic sample N=4803).

**Figure S1.** Flow chart of participants’ inclusion criteria for the final machine learning prediction model

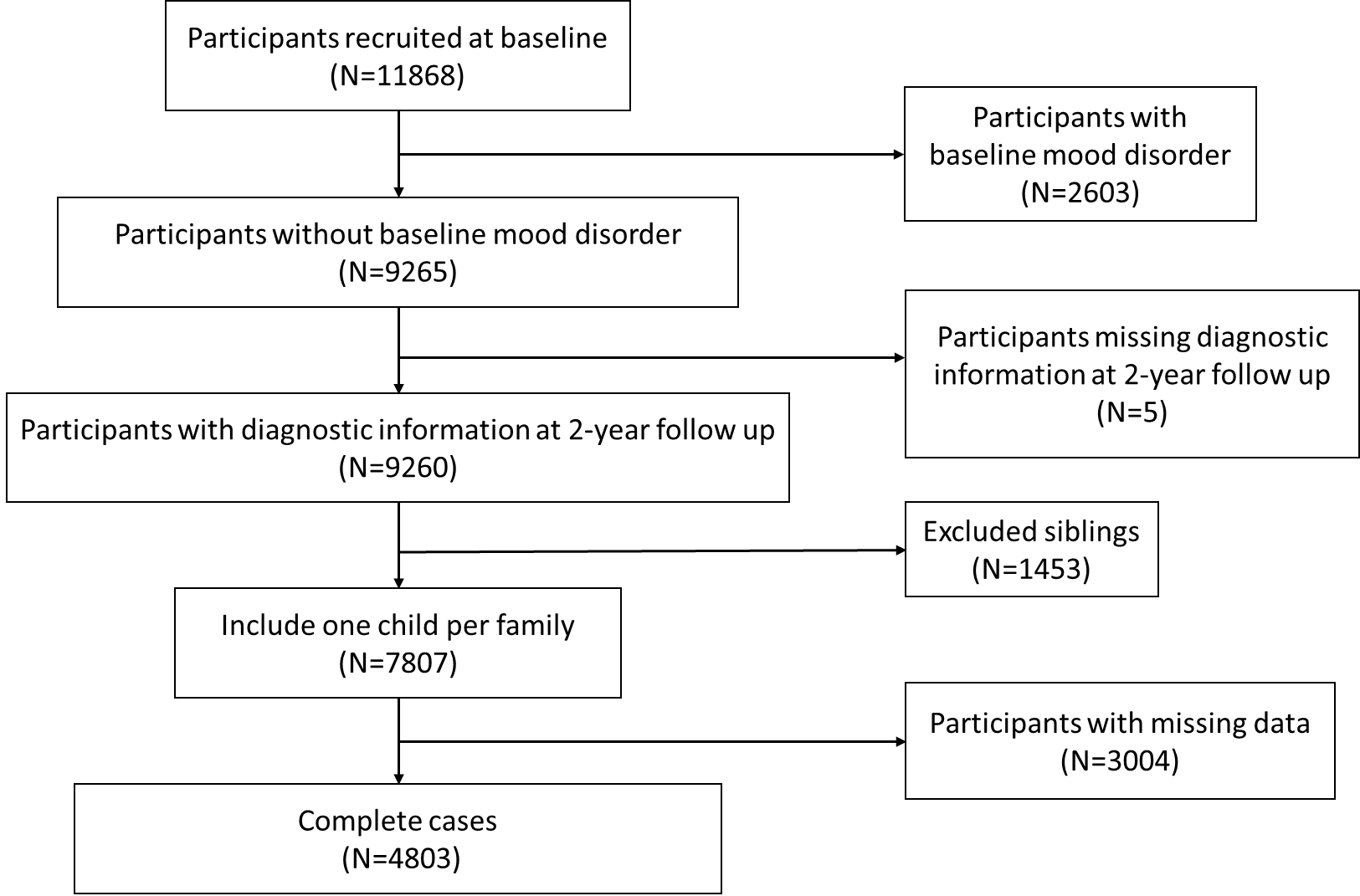

**Table S2.** Variable names and description of risk factors included at baseline, with respective assessments from which they were extracted

| **Variable names** | **Description** | **Measures** |
| --- | --- | --- |
| *Mental health* | | |
| anxiety | A composite for any anxiety diagnosis, present or past. | Kiddie Schedule for Affective Disorders and Schizophrenia (KSADS-5)^2–4^, parent report on their children. |
| eating_disorder | A composite for any eating disorder diagnosis, present or past. |  |
| ADHD_TIER2 | Attention Deficit Hyperactivity Disorder (ADHD) diagnosis, present or past. Participants were considered to have an ADHD diagnosis if (1) they had at least 6 inattentive and/or hyperactive-impulsive symptoms and impairment across two or more settings, (2) they had an estimated IQ over 70 (based on the Wechsler Intelligence Scale for Children – Fifth Edition (WISC-V) Matrix Reasoning total scale score > 3), and (3) they did not meet criteria for schizophrenia or bipolar disorder. |  |
| other_ASD | Other Specified Neurodevelopmental Disorder (Autism Spectrum Disorder). |  |
| psychotic | A composite for any psychotic diagnosis present or past. |  |
| disruptive_behavior | A composite for any disruptive behavior diagnosis present or past. |  |
| diag_com | A composite for any non-suicidal self-injury present or past. |  |
| cbcl_scr_syn_anxdep_r | Scores of parent reported syndrome scales (anxious/depressed; withdrawn/depressed; somatic complaints; social problems; thought problems; attention problems; rule-breaking behavior; and aggressive behavior.of child and adolescent mental health problems occurring in the past 6 months. | Child Behavior Checklist^5^, parent report on their children. |
| cbcl_scr_syn_withdep_r |  |  |
| cbcl_scr_syn_somatic_r |  |  |
| cbcl_scr_syn_social_r |  |  |
| cbcl_scr_syn_thought_r |  |  |
| cbcl_scr_syn_attention_r |  |  |
| cbcl_scr_syn_rulebreak_r |  |  |
| cbcl_scr_syn_aggressive_r |  |  |
| pps_y_ss_severity_score | Prodromal Psychosis Severity Score, measuring prodromal psychosis symptoms. | Prodromal Psychosis Scale^6–8^, child self-report. |
| pgbi_p_ss_score | Parent General Behavior Inventory - Mania, measuring subsyndromal mania. | Parent General Behavior Inventory^9^, parent report on their child. |
| sds_p_ss_total | Total score measuring sleep problems. | The Sleep Disturbance Scale for Children (SDSC)^10,11^, parent report on their child |
| su_caff_ss_sum_calc | Sum of five caffeine questions measuring caffeine use, type and quantity. | Caffeine Intake, child self-report |
| famhx_ss_parent_alc_p | Family History Assessment, measuring family history of psychopathology and substance use (for biological or adoptive parent). Includes: alcoholism, drug use, depression, trouble (holding a job, fighting, police interference), nervousness (nerves, nervous breakdowns) and visiting a doctor/counsellor for emotional/mental problem(s). | Family History Assessment^12^, parent self-report |
| famhx_ss_parent_dg_p |  |  |
| famhx_ss_parent_dprs_p |  |  |
| famhx_ss_parent_trb_p |  |  |
| famhx_ss_parent_nrv_p |  |  |
| famhx_ss_parent_prf_p |  |  |
| asr_scr_anxdep_r | Anxious/Depressed Adult Self Report (ASR) Syndrome Scale (raw score) measuring the levels of anxiety and depression of the participant (anxious/depressed; withdrawn/depressed; somatic complaints; social problems; thought problems; attention problems; rule-breaking behavior; aggressive behavior.) | Adult Self Report^13^, parent self-report. |
| asr_scr_withdrawn_r |  |  |
| asr_scr_somatic_r |  |  |
| asr_scr_thought_r |  |  |
| asr_scr_attention_r |  |  |
| asr_scr_aggressive_r |  |  |
| asr_scr_rulebreak_r |  |  |
| asr_scr_intrusive_r |  |  |
| *Cognition and dispositional traits* | | |
| nihtbx_picvocab_uncorrected | (National Institutes of Health) NIH Toolbox Picture Vocabulary Test (Ages 3+), measuring language: vocabulary knowledge, estimated verbal IQ. | NIH Toolbox Tasks^14^, child self-report |
| nihtbx_flanker_uncorrected | NIH Toolbox Flanker Inhibitory Control and Attention Test (Ages 8-11), measuring attention, cognitive control; executive function; inhibition of automatic response. |  |
| nihtbx_list_uncorrected | NIH Toolbox List Sorting Working Memory Test (Age 7+), measuring working memory, information processing. |  |
| nihtbx_cardsort_uncorrected | NIH Toolbox Dimensional Change Card Sort Test (Ages 8-11), measuring executive function: set shifting, flexible thinking; concept formation. |  |
| nihtbx_pattern_uncorrected | NIH Toolbox Pattern Comparison Processing Speed Test (Age 7+), measuring information processing; processing speed. |  |
| nihtbx_picture_uncorrected | NIH Toolbox Picture Sequence Memory Test (Age 8+), measuring episodic memory; sequencing. |  |
| nihtbx_reading_uncorrected | NIH Toolbox Oral Reading Recognition Test (Age 3+), measuring language: oral reading (decoding) skills, academic achievement. |  |
| cash_choice_task | Hypothetical question about whether the participant would prefer to receive a smaller amount of money immediately or a larger amount gradually. Measures delay aversion. | Cash Choice Task^15^, child self-report |
| ravlt_forget | The Rey Auditory Verbal Learning Test, used for assessing episodic memory | The Rey Auditory Verbal Learning Test^16^, child self-report |
| ravlt_learn |  |  |
| ravlt_immediate |  |  |
| ravlt_percent_forget |  |  |
| pea_wiscv_trs | Matrix Reasoning Total Raw Score, measuring the participant's fluid intelligence and visuospatial reasoning | WISC-V Matrix Reasoning subtest^17^, child self-report |
| upps_y_ss_negative_urgency | Urgency-Premeditation-Perseverance-Sensation Seeking-Positive Urgency (UPPS-P) for Children Short Form (ABCD-version), measuring impulsivity. Includes: negative urgency, lack of planning, sensation seeking, positive urgency and lack of perseverance. | UPPS-P for Children - Short Form (ABCD version)^18^, child self-report. |
| upps_y_ss_lack_of_planning |  |  |
| upps_y_ss_sensation_seeking |  |  |
| upps_y_ss_positive_urgency |  |  |
| bis_y_ss_bis_sum | Behavioral Inhibition/ Behavioral Approach System (BIS/BAS) Scales, measuring behavioral inhibition, and approach behaviors reward responsiveness, drive and fun seeking). | Behavioral Inhibition/ Behavioral Approach System Scales (BIS/BAS)^19^, child self-report. |
| bis_y_ss_bas_rr |  |  |
| bis_y_ss_bas_drive |  |  |
| bis_y_ss_bas_fs |  |  |
| *Demographic characteristics, environment, and culture* | | |
| demo_ed_v2 | Individual question about child's current school year/grade | Demographic questions^20^, parent report. |
| demo_prnt_empl_v2 | Individual question about current employment |  |
| demo_gender_id_v2 | Gender |  |
| highest_edu_p | highest education of parents |  |
| demo_prnt_marital_v2 | Individual question about participant's marital status. |  |
| accult_q1_y | Individual question about how well the participant speaks English. |  |
| accult_q2_y | Individual question about whether the participant speaks or understands a language/dialect other than English. |  |
| interview_age | Participant's age in months at baseline. |  |
| accult_q1_p | Individual question on proficiency of English speaking |  |
| accult_q2_p | Individual question on ability to speak/understand languages other than English |  |
| pmq_y_ss_mean | Parental monitoring mean, measuring parental monitoring or supervision. | Parental Monitoring Survey^21^, parent report. |
| fes_y_ss_fc_pr | Conflict Subscale from the Family Environment Scale (youth report). Measures family dynamics, cohesion, expressiveness, conflict. | Family Environment Scale - Family Conflict Subscale^22^, parent report and child self-report. |
| crpbi_y_ss_parent | Acceptance Subscale from Children’s Report of Parental Behavior Inventory (CRPBI). Measures environment - family & religion. | Children's reports of parental behavior Inventory (CRPBI)^23^, child self-report. |
| crpbi_y_ss_caregiver | Acceptance Subscale Mean of Report by Secondary Caregiver by youth |  |
| rules_drinking | Parental rules on drinking, smoking and marijuana use | Parent Rules^24^, parent report. |
| rules_smoking |  |  |
| rules_marijuana |  |  |
| srpf_y_ss_ses | School Risk & Protective Factors (SRPF) Survey, measuring risk and protective factors. Includes: School Environment Subscale. School Involvement Subscale & School Disengagement Subscale. | School Risk & Protective Factors Survey^25^, child self-report. |
| srpf_y_ss_iiss |  |  |
| srpf_y_ss_dfs |  |  |
| nsc_p_ss_mean_3_items | Neighborhood Safety Protocol, measuring risk and protective factors, crime. | Neighborhood Safety/Crime Survey^26^, parent report. |
| meim_p_ss_exp | Multi-Group Ethnic Identity - Revised (MEIM-R) Survey, measuring cultural affiliation. Includes: MEIM-R Exploration Subscale & MEIM-R Commitment and Attachment Subscale. | Multi-Group Ethnic Identity - Revised Survey (MEIM-R)^27^, parent report. |
| meim_p_ss_com |  |  |
| macv_p_ss_fs | Mexican American Cultural Values Scale , measuring familism, religion, independence, self-reliance. Includes: Family Support Subscale, Family Obligation Subscale, Independence & Self-Reliance Subscale, Family as Referent Subscale, and Religion Subscale | Mexican American Cultural Values Scale (MACVS)^28^, parent report. |
| macv_p_ss_fo |  |  |
| macv_p_ss_isr |  |  |
| macv_p_ss_fr |  |  |
| macv_p_ss_r |  |  |
| psb_p_ss_mean | Mean score of the Prosocial Behavior Subscale of Parent Report on Youth, measuring resilience | Prosocial Behaviour Subscale of Parent Report on Youth^29,30^, parent report on their child. |
| psb_y_ss_mean | Prosocial Behavior Subscale, measuring resilience, youth self report. | Prosocial Behavior Survey^29,30^, child self-report. |
| resiliency5a_reco | The number of friends (girls or boys, separately) and close friends (girls or boys, separately) were recoded as follows: 0 = 0; 1 = 1; … 10 = 10; 11–15 = 11; 16–20 = 12; 21–25 = 13; 26–30 = 14; and 31–100 = 15. | Youth Resilience Scale, child self-report. |
| resiliency5b_reco |  |  |
| resiliency6a_reco |  |  |
| resiliency6b_reco |  |  |
| weighted_average_passive_watching | Screen time for social media, gaming, and other activities, calculated as a weighted average across weekdays and weekends, reflecting the average daily use over the week. Passive watching referred to time spent watching television or movies; active gaming captured time spent playing video games on a computer or other devices; social engagement reflected time spent texting or video calling on a cellphone, tablet, or computer; and social media represented time spent on social networking platforms. | Screen Time Survey^31,32^, child self-report. |
| weighted_average_active_gaming |  |  |
| weighted_average_social_media |  |  |
| weighted_average_social_engagement |  |  |
| *Physical health* | | |
| anthro_waist_cm | Waist circumference (in inches). | Physical health - measured on site^33^ |
| BMI | Measurement of participants Body Mass Index (BMI) score |  |
| extracurricular | Frequency of extracurricular activities | Sports and Activities Involvement Questionnaire^34^, parent report |
| freq_sport | Frequency of sports activities |  |
| devhx_ss_8_alcohol_max_p | Individual question about maximum alcoholic drink intake in one sitting. | Developmental History Questionnaire^35^, parent report. |
| devhx_ss_8_alcohol_avg_p | Individual question about average alcoholic drink intake per week. |  |
| devhx_ss_12_p | Individual question about how premature (in weeks) the child was at time of birth. |  |
| medhx_ss_4b_p | Individual question about frequency of visits to emergency room in the past year | Medical History Questionnaire^36^, parent report. |
| medhx_ss_5b_p | Individual question about frequency of visits to emergency room before the past year |  |
| medhx_ss_6a_times_p | Individual question about frequency of broken bones |  |
| medhx_ss_6b_times_p | Individual question about frequency of sprains |  |
| medhx_ss_6c_times_p | Individual question about frequency of scrapes |  |
| medhx_ss_6d_times_p | Individual question about frequency of stitches |  |
| medhx_ss_6f_times_p | Individual question about frequency of falls |  |
| medhx_ss_6h_times_p | Individual question about frequency of high fever |  |
| medhx_ss_6i_times_p | Individual question about frequency of head injury |  |
| medhx_ss_9b_p | Individual question about frequency of undergoing general anaethesia or sedation for surgery or procedure |  |
| medhx_ss_6t_times_p | Individual question about frequency of other injuries |  |
| ehi_y_ss_score | Handedness score rating, measuring handedness; laterality quotient. | Edinburgh Handedness Inventory^37^, child self-report |

**Supplement 2.** Machine learning procedures

*Machine Learning Model Approach*

We implemented three supervised machine learning models to predict first-onset depression, using baseline risk factors as predictors and depression onset between baseline and a two-year follow-up as the binary outcome. The models were designed to capture the complex interplay of the risk factors while providing robust and reliable predictions. Our analysis focused on both the general population (first aim) and high-risk adolescents with ADHD (second aim).

*Data Preprocessing Pipeline*

The preprocessing pipeline was standardized across all three machine learning models to ensure consistency and avoid data leakage. The pipeline included several steps to prepare the data before fitting it into the models:

1. Standardization of Continuous Variables: All continuous variables were standardized (i.e., scaled to have a mean of zero and a standard deviation of one) to ensure they were on the same scale. This step is essential to avoid issues where variables with larger scales dominate the model, which is particularly important for algorithms like SVM and Elastic Net, which are sensitive to the magnitude of the input features.

2. Dummy Coding of Categorical Variables: Categorical variables with more than two levels were dummy-coded to convert them into numerical form. Variables with only two levels were left unchanged, as they already fit into a binary encoding suitable for the models.

*Model-Specific Hyperparameter Tuning*

For each of the three models, we incorporated hyperparameter tuning to enhance performance and optimise model fitting. The hyperparameters were selected based on the characteristics of each model and were optimized using grid search based on cross-validated AUC scores

For the SVM, we tuned the cost parameter (C) and the kernel type (linear and Radial Basis Function (RBF) kernels). This enabled us to control the degree of regularization performed by the model, while also testing the performance of linear and non-linear kernels.

The Elastic Net regularization technique was applied, which combines both L1 (Lasso) and L2 (Ridge) penalties. This model requires the tuning of two main hyperparameters: the regularization strength (C) and the balance between L1 and L2 regularization (l1_ratio).

For Random Forest, hyperparameters such as the number of trees (n_estimators), maximum tree depth (max_depth), and minimum samples per leaf (min_samples_leaf) were fine-tuned. These hyperparameters determine the complexity of the model, and allowed us to identify a model that was sensitive to patterns within the data without overfitting the data.

*Feature Selection and Near-Zero Variance Filtering*

To prevent the inclusion of irrelevant or redundant features, we implemented a near-zero variance filter. This filter was applied to the predictors before the model fitting process. Features with extremely low variance (i.e., those that do not vary much across the data) were removed, as they do not contribute meaningfully to the model’s predictive power. This filtering was applied to each model separately, and the threshold value was optimized as a hyperparameter, with values of: 0.01, 0.03, 0.05, and 0.1 tried.

*Model Evaluation*

The evaluation metrics included accuracy, precision, recall, and F1-score, and confusion matrices were generated to assess classification performance. We also used ROC curves and AUC scores to compare model discrimination across different thresholds.

In summary, the pipeline was designed to preprocess and tune the models systematically, ensuring that all models received the same preprocessing steps and that hyperparameters were optimized for each. The result was a robust and well-calibrated machine learning model that could predict first-onset depression based on baseline risk factors.

**Supplement 3.** Imbalanced dataset solution

Our outcome variable was highly imbalanced, with approximately 10% of participants developing first-onset depression at the two-year follow-up. This group was of primary interest, making the class imbalance a critical issue for model performance. Class imbalance poses significant challenges for machine learning models, which often assume balanced class distributions and may otherwise be biased toward the majority class ^38,39^. To solve this issue, we implemented two commonly used strategies: resampling techniques and algorithm-level adjustments.

First, we tested synthetic minority over-sampling using SMOTE (Synthetic Minority Oversampling Technique), which generates synthetic examples of the minority class to improve the training distribution^40,41^. Second, we incorporated class weighting into the model training process, which assigns higher misclassification costs to the minority class, encouraging the model to prioritize correct classification of depressed cases. Most machine learning libraries, including scikit-learn, allow for class weights to be set either manually or automatically (class_weight = "balanced"), based on the inverse class frequencies.

We systematically compared these strategies across all models. While both approaches improved the models’ ability to identify the minority class, using class weights alone yielded better predictive performance. As such, we selected the class-weighted models for our final results.

**Table S3**. Confusion matrices for the performance of machine learning models.

|  | True positive | True Negative | False Positive | False Negative | Sensitivity | Specificity | PPV | NPV |
| --- | --- | --- | --- | --- | --- | --- | --- | --- |
| Full sample | | | | | | | | |
| Elastic Net | 57 | 618 | 248 | 38 | 0.60 | 0.71 | 0.19 | 0.94 |
| Random Forest | 29 | 763 | 103 | 66 | 0.31 | 0.88 | 0.22 | 0.92 |
| SVM | 46 | 702 | 164 | 49 | 0.48 | 0.81 | 0.22 | 0.94 |
| Cross sample | | | | | | | | |
| Elastic Net | 19 | 38 | 60 | 9 | 0.68 | 0.39 | 0.24 | 0.81 |
| Random Forest | 16 | 62 | 36 | 12 | 0.57 | 0.63 | 0.31 | 0.84 |
| SVM | 21 | 44 | 54 | 7 | 0.75 | 0.45 | 0.28 | 0.86 |
| ADHD sample | | | | | | | | |
| Elastic Net | 8 | 62 | 33 | 14 | 0.36 | 0.65 | 0.20 | 0.82 |
| Random Forst | 2 | 89 | 6 | 20 | 0.09 | 0.94 | 0.25 | 0.82 |
| SVM | 8 | 67 | 28 | 14 | 0.36 | 0.71 | 0.22 | 0.83 |

Abbreviations: SVM = Support Vector Machine, PPV = Precision/Positive predictive Value, NPV = Negative Predictive Value.

**Figure S2** ROC curve for the streamlined prediction model (top 10 most important features) in the whole sample.

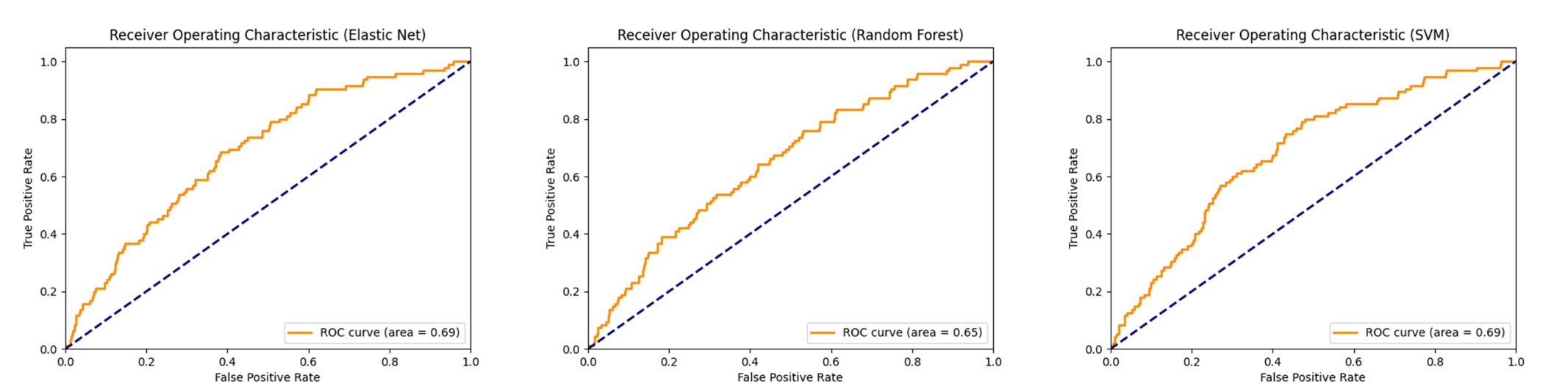

Abbreviations: ROC = Receiver Operating Characteristic. SVM = Support Vector Machine
